# Timely Geographic Access to Blood Banking Facilities: A Pan-India Modeling Study

**DOI:** 10.1101/2025.05.18.25327878

**Authors:** Gaurav Urs, Aamir Miyajiwala, Ankit Raj, Siddhesh Zadey

## Abstract

**Background:** Efficient blood transfusion planning requires knowledge of the distribution and accessibility of blood banking facilities (BBFs). We mapped population-level, timely geographic access to BBFs across 729 districts across India.

**Study Design and Methods:** We conducted a pan-India geospatial modeling study using 2024 data from e-RaktKosh, a centralized blood management system. As outcomes, we estimated BBFs per million people, travel time to the nearest BBF, and access population coverage (APC), defined as the proportion of the population with access to the nearest BBF by motorized transport (60 minutes) and walking (30 minutes).

**Results:** In 2024, India had 5685 BBFs, averaging 4.16 facilities per million people. Median travel times to the nearest BBF were 35.25 minutes by motorized transport and 253.27 minutes by walking. For APCs, 93.86% of people were within 60 minutes of their nearest BBF by motorized transport, while 16.21% were within 30 minutes by walking. There were large differences in APCs across districts and states, with notable rural-urban disparities.

**Discussion:** Access to blood banking facilities in India exhibits geographic disparities, with the northeastern and island Union Territories with prolonged median travel times and low APC values. Rural areas are disproportionately adversely affected. Future efforts should focus on using timely geographic access data to inform strategic infrastructure planning and resource allocation, enhancing healthcare equity.

## 1 Introduction

The World Health Organization (WHO) classifies blood and blood products as essential medicines and notes a significant gap in access to these critical resources across countries, adversely impacting access to blood in low- and middle-income countries.^1^ Timely geographic access to blood and blood products is essential across acute specialties, including emergency, intensive care, surgical, obstetric, trauma, and anesthesia care. Beyond acute specialties, such access is vital for managing chronic hemato-oncological conditions that require regular transfusions.^2^ Hence, establishing efficient and equitable blood banking systems is arguably the most important way to meet the above-mentioned healthcare needs.^3^

Timely geographic access to blood relies, partly, on spatial proximity to BBFs. Prolonged travel times delay access to blood transfusions, potentially increasing the risk of adverse outcomes, especially for patients with critical or chronic conditions or who need emergent or urgent care.^4^ Hence, access time to BBFs is vital for the equitable utilization of blood banking services.^5^

In low- and middle-income countries (LMICs), the blood supply relies primarily on blood banks and storage facilities. LMICs are known to face barriers concerning availability, safety, and infrastructure.^5,6^ While studies in LMICs have focused on these challenges, efforts to evaluate timely geographic access to BBFs are limited and localized to smaller geographies, regardless of the high priority.^7,8^

India, the most populous country with over 1.4 billion residents, faces formidable challenges in ensuring access to a large and heterogeneous blood banking system regulated under the National Blood Transfusion Council (NBTC).^9^ Previously, studies have evaluated the performance of Indian BBFs for blood volume availability, accreditation, safety, infrastructure, regulations, etc., using systematic composite indices.^10,11^ However, they did not include timely geographic access, which is the primary focus of this study. Recent regional studies have identified areas for focus as ‘Blood Deserts’, and building on these findings, we aim to provide a pan-Indian assessment with resolution at the district level and an urban-rural divide. ^12^

We aimed to assess the various measures of geographic access to BBFs in India. First, we mapped the BBF geolocations and estimated BBF density per million people. Second, we analyzed the travel times to the nearest BBFs by different modes of transport. Third, we assessed the proportion of people who can access BBFs within 30 and 60-minute intervals, which we defined as population access coverage (APC). We estimated the above-mentioned outcomes of geographic accessibility at national, state, and district levels with rural and urban disaggregations. Finally, we assessed state-level correlations between APC and previously published BBF performance indices as a step toward concurrent validation.

## 2 Methods

### 2.1 Data sources

We obtained geographic locations for 5685 blood bank centers and facilities (BBF) among Government, Red Cross, Charitable, and Private centers across 729 districts, within 28 states and eight union territories (UTs) from e-RaktKosh, a centralized blood management system maintained by the Centre for Development of Advanced Computing (CDAC).^13^

All available records across states and union territories were manually extracted into Microsoft Excel and reviewed for duplicate entries and omissions to ensure accuracy. Variables included state, facility name, address, latitude, longitude, contact number, email address, and ownership category, with geographic coordinates. Facilities under e-RaktKosh are classified into hospital-based blood banks, standalone blood centers, blood storage units (BSUs) attached to secondary/rural health facilities, and institutional blood centers. The extracted centers were collectively labeled as “Blood Bank Facilities” for the purpose of the study.

We used the Awesome Table API to geocode the locations. Coordinate plots of latitude versus longitude were used to identify geographic outliers deviating from India’s expected coordinate cluster. Facilities with coordinates outside India’s boundaries (6°N–37°N latitude, 68°E–98°E longitude) were flagged and manually verified in Google Maps, and their addresses were corrected.

Population counts were sourced from high-resolution (1 km^2^) United Nations-adjusted projections for 2020 from the WorldPop dataset.^14^ We obtained friction surface 2019 rasters for motorized transport and walking from the Malaria Atlas Project (MAP), an international scientific effort, primarily concerned with mapping the global response to malaria.^15^ We used these rasters to calculate travel times to BBF from each square kilometer (1 km²) to BBFs.^16^ MAP and WorldPop are highly cited global sources that have been previously verified.^17,18^

We obtained the administrative boundaries of India from publicly available and validated shapefiles.^19^ A previously published global raster with ordinal catchment area (CA) categories based on population densities and proximity to high-density urban areas provides standardized rural-urban regions. We classified CA categories >7 as rural areas. This threshold corresponds to the URCA classification methodology, where categories 1–7 represent the seven urban agglomeration sizes based on population (ranging from large cities with >5 million inhabitants to towns of 20,000–50,000 people), while categories 8–30 represent rural catchment areas classified by travel time (<1 hour, 1–2 hours, 2–3 hours) to these urban centers and hinterland regions beyond 3 hours from any urban agglomeration of at least 20,000 people.

The data sources used in this study range over the period of 2019 to 2024 (e-RaktKosh (2024), WorldPop (2020), and MAP (2019)). Although these datasets span over 5 years, the blood bank locations, road infrastructure, and population spatial distributions change gradually over time.

### 2.2 Outcomes

We studied three outcomes. First, BBF density was examined as the number of BBFs per million people to understand the geographic distribution of BBFs relative to the underlying population residing in a given geography. Second, the median travel time to the nearest BBF was investigated. Travel time estimates for each 1 km² raster were used, and the median and interquartile range (IQR) were reported at the aggregate group level. Finally, the access population coverage (APC) was defined as the percentage of the population with timely access to the nearest BBF. We used 60 minutes as the timely access threshold for travel by motorized transport and 30 minutes for walking. Travel time thresholds such as 30 minutes for walking and 60 minutes for motorized travel are commonly used in spatial accessibility research to reflect meaningful bounds on how far individuals will travel for health services^32^. For example, survey evidence shows that most adults report current travel to primary care is ≤30 minutes, and only a small proportion exceeds 60 minutes, with average acceptable travel times clustering around these values for both urban and rural residents. This supports the use of 30 minutes and 60 minutes as pragmatically relevant cut-offs for walking and motorized accessibility measures in health access analyses.The APC integrated both the population and timeliness aspects of access.

### 2.3 Analysis

We manually cleaned the BBF addresses to improve machine readability before geocoding, performed using the Google Maps application programming interface (API) and the ‘Awesome Table’ add-on for Google Sheets. Any BBFs not captured were manually geocoded in Google Maps. The locations with the most relevant address strings were selected from those that returned multiple sets of coordinates. Geo-coordinate similarity was used to identify duplicate facilities using exact coordinate matching (rounded to 4 decimal places, ∼11m precision) combined with fuzzy name matching (>90% string similarity) for facilities within 1 km proximity. Points beyond India’s geographic extent (latitude 6°N–37°N, longitude 68°E–98°E) were excluded.

The WorldPop 2020 and rural-urban CA rasters were analyzed to estimate population distribution at various geographic levels. The global multi-category CA raster (1 km² resolution at the equator, with each pixel representing an agglomeration category) was first cropped to fit within India’s national boundaries. The raster was then divided into two layers: rural (CA agglomeration categories >7) and urban (CA agglomeration categories ≤7). We then overlaid the WorldPop 2020 population raster for India (also 1 km² resolution) on these binarized rural-urban CA rasters to get rural and urban population rasters. To ensure compatibility, the CA raster was resampled to match the population raster’s extent, resolution, and origin. In the rural population raster, rural area pixels were assigned a value of 1, while urban areas were marked as ‘NA’, and vice versa in the urban population raster. Residential population in each pixel was calculated by multiplying the population count by the corresponding category value (1 for rural areas in the rural raster). The total population was derived by summing the rural and urban populations. This was done at all geographic levels: subnational (districts and states) and national (all-India). It is important to note that not all districts had both rural and urban areas, as some were entirely rural. In this analysis, 728 districts had rural areas, while 691 districts had urban areas.

We calculated the travel times based on the shortest time taken to traverse the MAP friction surface from each 1 km² pixel to its nearest BBF location.^18^ Two accessibility rasters were generated based on the modes of transport: one for walking and the other for motorized transport, providing travel times in minutes assuming optimal speeds. We estimated the median and interquartile range (IQR) for travel times across the district, state, and national levels.

For APC calculations, we used timely-access thresholds (60 minutes for motorized transport and 30 minutes for walking) to create binary accessibility rasters, in which pixels below the threshold were coded 1 and those beyond were marked as ‘NA’. The accessibility raster was then aligned and overlaid onto the population raster to determine the population with timely access. We calculated the sum of pixels coded ‘1’ to get the population with timely access at different geographic levels, i.e., district, state, and national levels. Finally, the proportion of the population with timely access (i.e., the APC) was calculated by dividing the population with access in a region by its residential population for rural and urban disaggregations.

We conducted the above geospatial analyses in R software (version 4.3.2). Overall, they are similar to our recent work on population-level timely geographic access to medical college hospitals, COVID testing centers, primary and community healthcare centers, surgical care facilities, and palliative care centers in India.^20–24^

Previously, we analyzed blood banking performance at the state and district levels using 2016 NBTC data.^11^ In that analysis, we constructed a composite index made up of 49 state-level variables across seven themes, including accreditation, ownership, regulation, infrastructure, safety, volume, and workforce. As an initial concurrent validation (i.e., sanity check) for the timely access work presented here, we assessed the ecological relationships between the APC values for BBFs and the previously described BB performance index using Spearman correlations. We conducted the analysis using the SR Plot tool. We classified the strength of correlation as follows: very strong (0.7 to 0.9), strong (0.4 to 0.7), moderate (0.3 to 0.4), weak (0.2 to 0.3), and negligible (0.1 to 0.2), and used the conventional 5% alpha threshold for determining statistical significance.^26^ We used state-level data rather than district-level data for this analysis because district boundaries have undergone significant changes between 2016 and 2024.

## 3 Results

### 3.1 Density of BBFs

In 2024, India had 5685 BBFs. Among states/UT, Tamil Nadu had the highest number of BBFs (642), while the UT of Lakshadweep had just one BBF **(Figure 1A)**. The national BBF density was 4.16 facilities per million people **(Table 1)**. The BBF density ranged from 1.34 in Bihar to 12.94 in Lakshadweep. Thirteen (36%) states had BBF density below the national value. Among 729 districts, Bangalore in Karnataka had the highest number of BBFs (104), while 108 districts had only one BBF each. Regarding BBF density, Serchhip in Mizoram had the highest BBF density (40.54), while Suburban Mumbai (n=56) in Maharashtra had the lowest (0.19) (**Figure 1B)**. Three hundred and sixty-four districts had BBF density lower than the national value. Generally, districts in Tamil Nadu, Kerala, and South Karnataka had higher densities, while those in northern and eastern states of Bihar, Uttar Pradesh, West Bengal, and Jharkhand had lower BBF densities.

**Figure 1:**
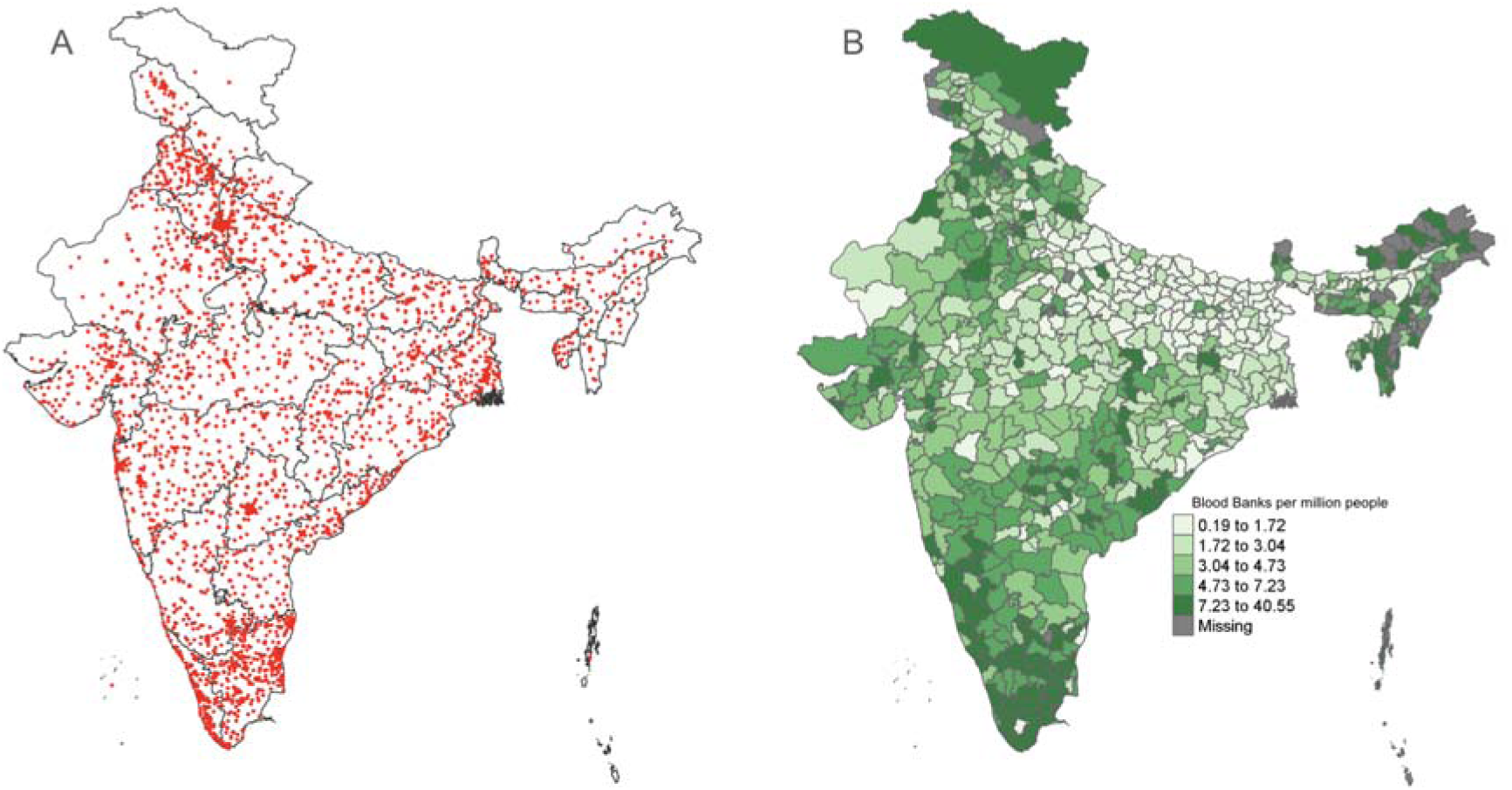
A) Geographic locations and B) density per million people of blood banking facilities across 729 districts.

**Table 1:** Summary of national, urban, and rural values for median access times and access population coverage.

| Value | National | Urban | Rural |
| --- | --- | --- | --- |
| Median (Interquartile Range) motorized travel time in minutes | 35.25<br>(20.20–62.40) | 6.66<br>(2.37–20.22) | 35.91<br>(20.99–63.05) |
| Median (Interquartile Range) walking travel time in minutes | 253.27<br>(153.11–392.3) | 66.65<br>(25.83–188.33) | 256.59<br>(157.87–394.67) |
| Access population coverage (APC) within motorized transport by 60 minutes | 93.86% | 99.44% | 91.83% |
| Access population coverage (APC) within walk by 30 minutes | 16.21% | 53.02% | 2.63% |

### 3.2 Travel time to the nearest BBF

#### 3.2.1 Motorized Transport

By motorized transport, the national median travel time to the nearest BBF was 35.25 minutes (IQR: 20.20–62.40). For urban areas, the median travel time was 6.66 minutes (IQR: 2.37–20.22), about 5 times lower than that for rural areas: 35.91 minutes (IQR: 20.99–63.05) **(Figure 2A)**. Among states/UTs, Lakshadweep had the shortest median travel time to the nearest BBF at 1.40 minutes (IQR: 0.70–1.77), which was over 25 times faster than the national median. Ladakh had the longest median travel time at 1781.16 minutes (IQR: 1125.50–2342.05). Across districts, Mumbai in Maharashtra recorded the shortest median travel time of 0.92 minutes (IQR: 0.52–1.35), about 38 times faster than the national average. Anjaw in Arunachal Pradesh had the longest median travel time of 1434.51 minutes (IQR: 991.29–1913.09), more than 40 times the national value.

**Figure 2:**
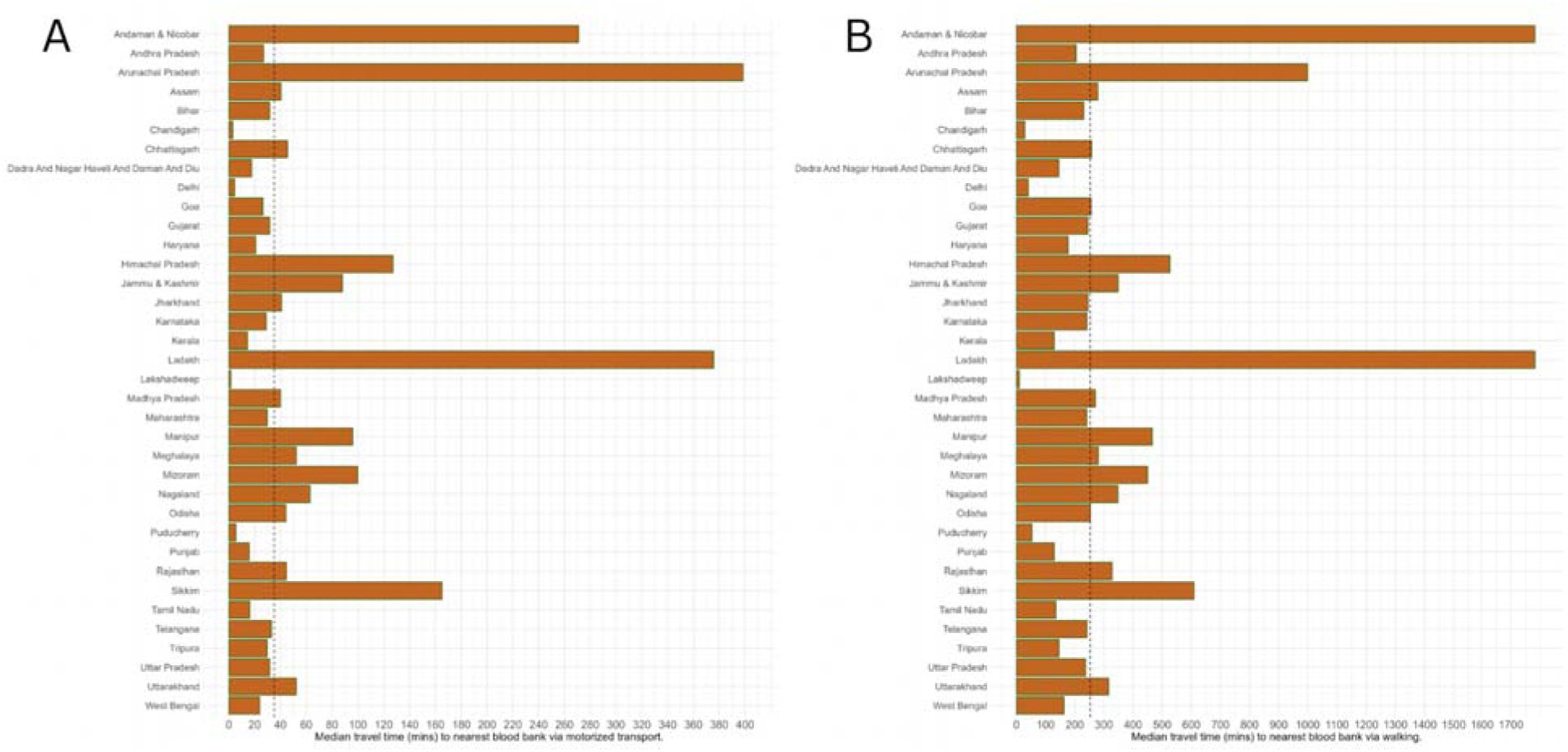
Median travel time (in minutes) to the nearest blood banking facility by A) motorized transport and B) walking across 36 states/Union Territories. The dotted lines note the national values.

#### 3.2.2 Walking

By walking, the national median travel time to the nearest BBF was 253.27 minutes (IQR: 153.11–392.38). For urban areas, the median travel time was 66.65 minutes (IQR: 25.83–188.33), about 4 times faster than that for rural areas: 256.59 minutes (IQR: 157.87–394.67) **(Figure 2B)**. Among states/UTs, Lakshadweep had the shortest median travel time to a BBF by walking, at 9.63 minutes (IQR: 4.81–13.01), over 26 times faster than the national value. Across districts, Lakshadweep in the Lakshadweep Islands recorded the shortest median travel time at 9.63 minutes (IQR: 4.81–13.01), over 26 times faster than the national median time. The North & Middle Andaman region of the Andaman and Nicobar Islands had the longest median travel time of 2331.40 minutes (IQR: 1617.12–2995.20), more than 9 times the national value.

### 3.3 Access Population Coverage (APC)

#### 3.3.1 Motorized Transport

Nationally, 1,291,542,656 people (93.86% of the Indian population) had access to BBFs within 60 minutes by motorized transport. Among the states/UTs, New Delhi and Chandigarh had the highest APC of 100%, while the Andaman and Nicobar Islands had the lowest APC of 48.56% **(Figure 3A)**. Forty-nine districts had an APC value of 100%, while three districts had an APC of 0% **(Figure 3B)**.

**Figure 3:**
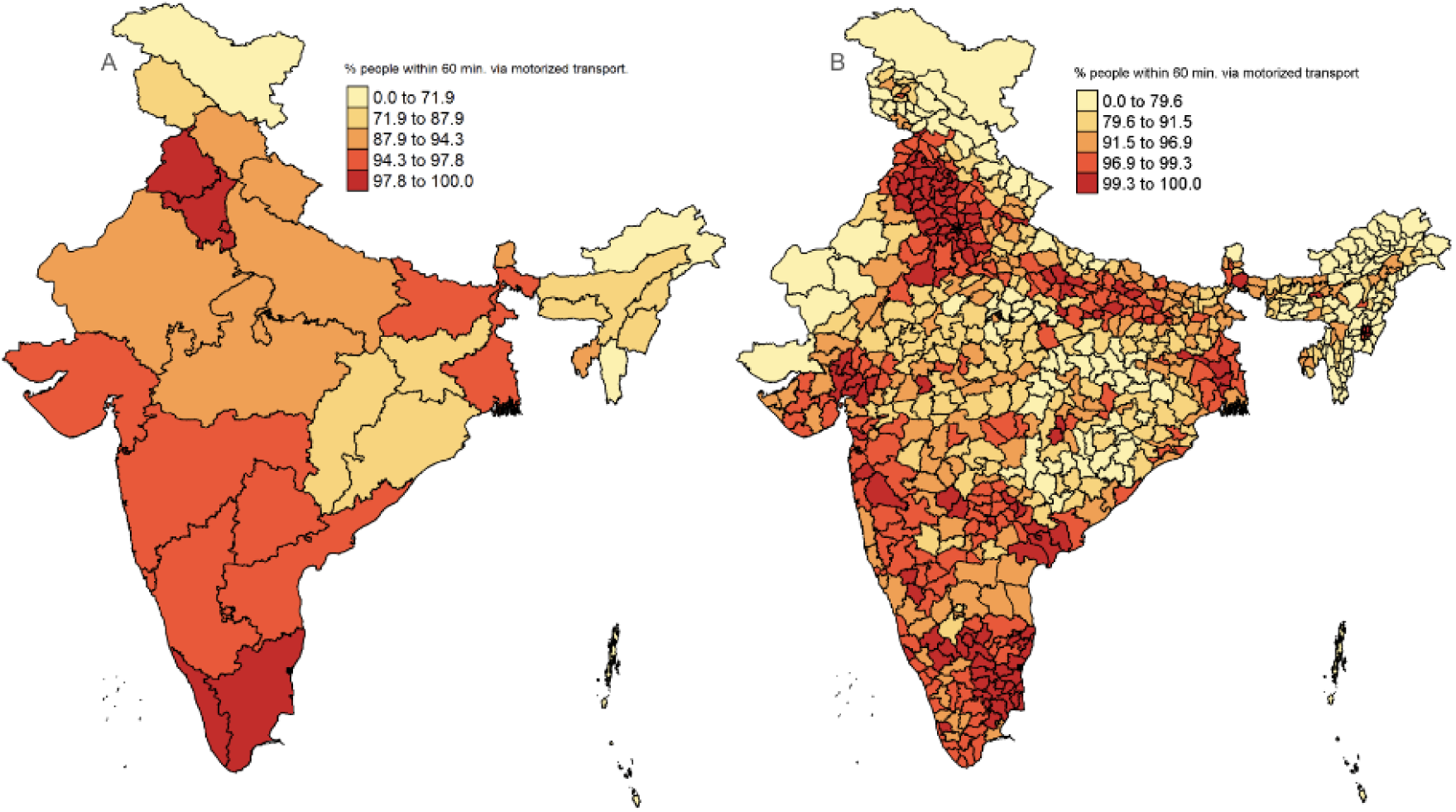
Access population coverage within 60 minutes by motorized transport for A) states/Union Territories and B) districts.

Nationally, 99.44% of the population residing in urban areas and 91.83% of those in rural areas could access their nearest BBF within 60 minutes. Urban areas of five states/UTs had an APC of 100%, while only two rural areas achieved the same. At the district level, 396 urban and 45 rural areas had 100% APC, while three rural and two urban areas had an APC value of 0% **(Figure 4)**.

**Figure 4:**
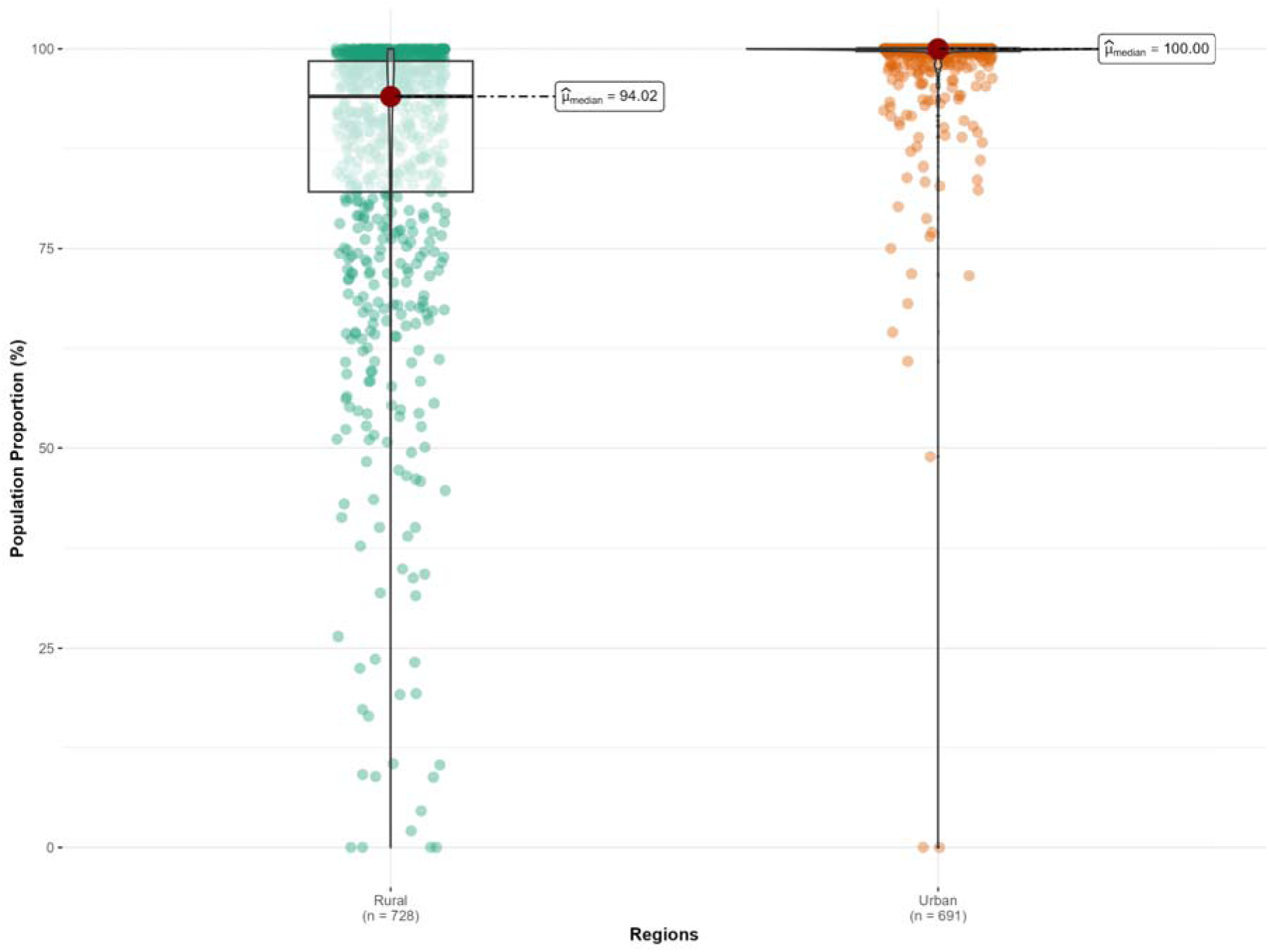
Access population coverage within 60 minutes by motorized transport in rural and urban areas across districts.

#### 3.3.2 Walking

Nationally, 223,158,576 people (16.21% of the Indian population) had access to BBFs within 30 minutes by walking. Among the states/union territories, New Delhi led with an APC of 69.05%, while Ladakh had the lowest value at 2.76% **(Figure 5A)**. Among districts, Shahdara, New Delhi had an APC of 97.94%, while 78 districts had an APC value of 0% **(Figure 5B)**.

**Figure 5:**
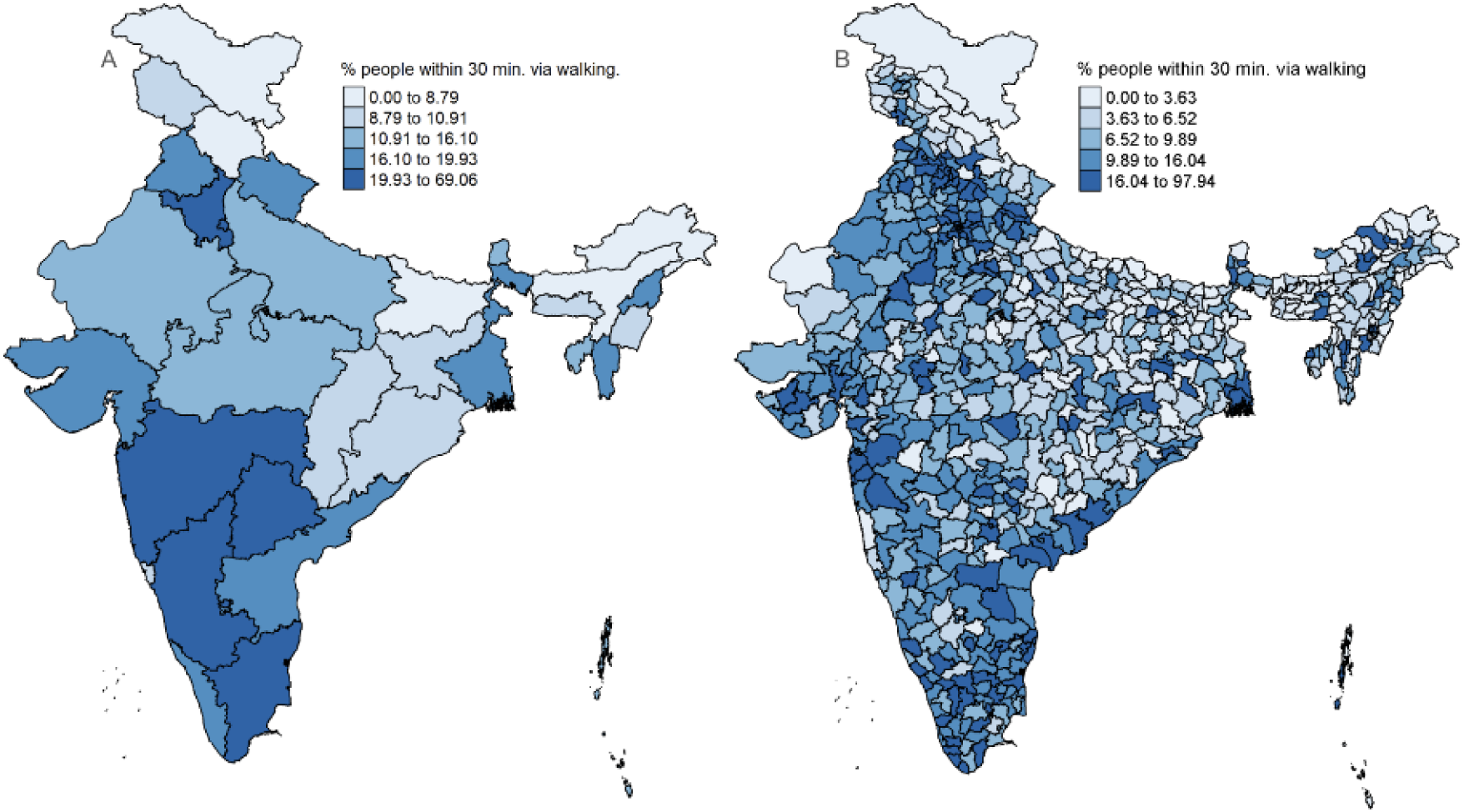
Access population coverage within 30 minutes by walking across A) states/Union Territories and B) districts.

Nationally, 53.02% of the population residing in urban areas and only 2.63% of rural areas had access to BBFs within 30 minutes by walking. Among states/UTs, rural Puducherry had the highest APC at 13.52%, while rural areas in two states/UTs had an APC of 0%. Urban areas in Mizoram recorded the highest APC of 91.76% while those in Bihar noted the lowest APC of 21.90%. At the district level, only the urban Lower Subansiri in Arunachal Pradesh had an APC of 100%, while urban areas in 30 districts had an APC of 0%. There were 48 rural areas across districts without access to BBFs within 30 minutes, while three areas had 100% APC **(Figure 6)**.

**Figure 6:**
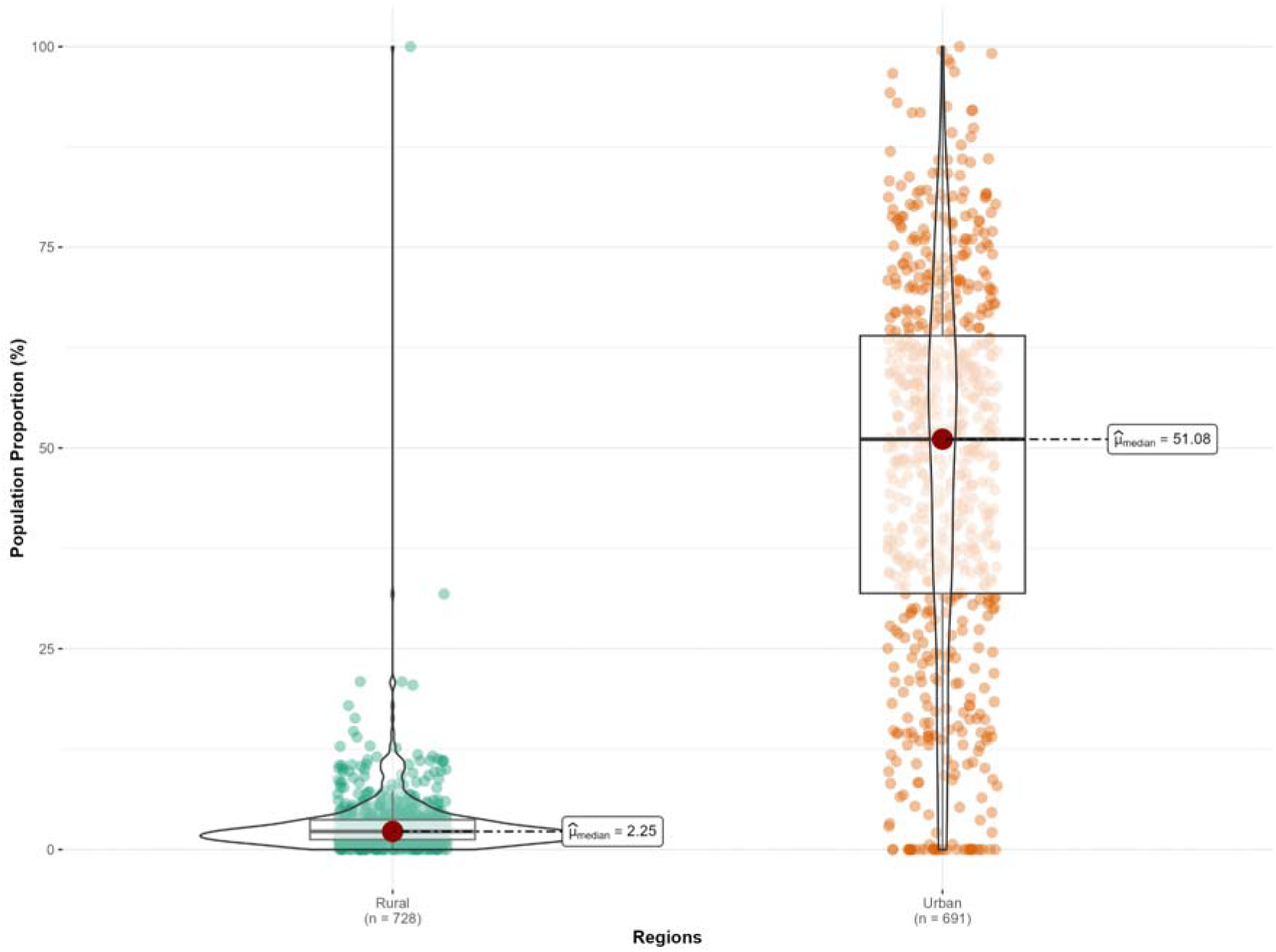
Access population coverage within 30 minutes by walking in rural and urban areas across districts.

### 3.4 Correlations with Blood Banking Performance Index

In a sample of 28 states and eight UTs, we found a moderate positive correlation between the blood banking performance index and APC values for APC within 60 minutes by motorized transport (R = 0.39, 95% CI 0.06–0.64; p = 0.022) and a strong positive correlation for APC within 30 minutes by walking (R = 0.42, 95% CI 0.11–0.66; p = 0.011). Both these were statistically significant at the alpha threshold of 0.05.

## 4 Discussion

### 4.1 Summary of findings

In 2024, India had 5,685 blood banking facilities (BBFs) serving about 1.4 billion people. The national density was 4.16 BBFs per million people, ranging from 1.34 to 12.94 BBFs per million people across states/UTs. The median walking time to the nearest BBF was 253.27 minutes, while the median time for motorized transport was 35.25 minutes. Nationally, 93.86% of the population could reach their nearest BBF in 60 minutes by motorized transport, and 16.21% could reach it within 30 minutes by walking. This meant that 88,521,598 people did not have access to a BBF within 60 minutes by motorized transport and 1,208,079,465 did not have access to the nearest BBF within 30 minutes by walking. Urban areas had almost universal access, whereas rural areas had lower coverage. We also observed heterogeneities in access outcomes across states/UTs and districts. At the state level, the APCs were correlated with the blood banking performance index, providing preliminary evidence of concurrent validity.

### 4.2 Previous studies and relevance of findings

In this study, we measured travel times over distances, as the duration to reach a facility depends on various factors beyond distance. Further, APC integrates population with travel time to offer a more comprehensive view of geographical accessibility to care. Previously, another modeling study, which focused more on blood availability, also assessed timely access in specific regions of the northern and northeastern Indian states.^12^ However, a pan-India assessment providing district-level values with rural-urban disaggregations was missing.

Our study noted disparities between rural and urban areas. Rural areas had lower BBF densities, longer travel times, and lower APC values than urban areas in India. These findings echo the observations of the Association of Rural Surgeons of India (ARSI) that highlighted the disparity in blood supply distribution between rural and urban areas, noting that while over two-thirds of the population resided in rural areas, most blood banks were concentrated in urban centers.^5^

### 4.3 Policy implications

Our findings help determine whether the lack of access to BBFs is due to a shortage of BBFs, challenges in reaching them, or both.

We noted disparities between rural and urban areas. Rural areas with low BBF density, such as Bihar, West Bengal, Uttar Pradesh, and Jharkhand, should prioritize establishing new BBFs to meet population demands. Several districts within these states have both low APC and prolonged travel times, indicating compounded geographical issues. Rural regions of Arunachal Pradesh, Assam, and parts of Uttar Pradesh face challenges in travel times mainly due to challenging terrain and poor road infrastructure. In these areas, deploying mobile blood banks and establishing satellite blood banks can help expand services and increase blood collection coverage. Furthermore, partnerships with non-governmental organizations and local community health workers can bridge coverage gaps.^27^

In urban areas, despite having a higher number of BBFs, access remains limited due to high population density and unequal distribution of blood banks, which are often concentrated in centers.^28^ This makes it difficult for people in peripheral regions to access blood banks during emergencies, especially in metropolitan cities such as Mumbai, New Delhi, and Kolkata.^29^ Districts, such as Suburban Mumbai, that have low BBF density due to high population denominators, highlight areas of priority where redistribution and decentralization of BBFs are effective solutions. Moreover, the public-private divide, the cost of blood and blood products, and emergency transport delays due to traffic congestion can further exacerbate the lack of timely access.^20^ To address these challenges, decentralizing blood bank functions, promoting real-time inventory systems (e.g., e-RaktKosh), establishing blood transport services, and educating the public about access points can help improve access population coverage.^13,30^

In states with longer travel times and where building BBFs might be challenging, such as Ladakh, Arunachal Pradesh, and Himachal Pradesh, the focus should shift to alternative solutions like walking blood banks, immediate autologous transfusion, and donor blood drop strategies.^27^ Rigorous mapping of ‘blood deserts’, i.e., regions lacking access to blood and blood products and associated healthcare services, can help inform local policy changes and implementation strategies.^31^

### 4.4 Strengths and limitations

To evaluate timely geographic access to BBFs in India, we employed a novel method using multiple outcome measures. Our pan-India study comprehensively analyzed the densities of BBFs per million people, travel times to the nearest BBF by two modes of transport (walking and motorized), and access population coverage, which combines population and timeliness measures, for 5685 BBFs across 729 districts. A novel feature of the study is the assessment of access across rural and urban areas. We also contribute to the broader global health data inventory by adding geolocation data for the Indian BBFs. The study focuses on timely geographic access beyond the common blood donation index, which is critical from a health systems perspective. These study methods can be adopted for similar analyses in other low-and middle-income countries.

Our study has several limitations. First, our analysis is subject to the assumptions and limitations of the original data sources, including e-RaktKosh, Worldpop, and MAP. WorldPop estimates are derived from census redistribution models and may not fully capture local population heterogeneity, recent demographic change, or short-term mobility. MAP travel-time rasters are based on cost-surface modeling with assumed travel speeds and do not reflect real-world variability in transport availability, road conditions, or health-system constraints. As a result, accessibility estimates should be interpreted as indicative and comparative rather than precise measures of realized access. Second, the APC estimates do not consider whether individuals have access to or own motor vehicles. Third, in regions of high population density, the low per-capita BBF density primarily reflects the large population denominator. Median travel times in regions with limited ground transport infrastructure likely represent upper-bound estimates due to the above limitations. Hence, these ratios should not be interpreted as inequity in isolation. Another factor to note is that of traffic congestion which can delay timely access. Fourth, our study calculates travel times and APCs based on walking and motorized land transport, without factoring in other modes of travel, such as waterways and airways, since travel time raster data to consider such complex travel scenarios remains unavailable. We interpret walking-based travel times as upper bounds and motorized travel times as lower bounds on accessibility, with true travel times likely lying between these two estimates. Fifth, we did not examine differences in access based on demographic factors such as age, gender, or socioeconomic status. Sixth, the policy implications suggested in this study are presented as potential strategies particular to the mode of transport, based on previous similar studies. We have not evaluated the feasibility, cost-effectiveness, regulatory compliance, and operational requirements of these suggested implications. Finally, the correlations between the blood banking performance index and APC values are exploratory since the data for these variables are collected at different time points. The analysis is ecological and can have aggregation bias. Even so, these findings are novel and pave the way for future studies that provide a more nuanced and comprehensive picture of geographical access to BBFs

## 5 Conclusion

In this study, we analyzed geographic access to blood bank facilities using multiple outcome measures, including population, travel times, and a combination of the two. Our study provides the first comprehensive pan-India assessment of timely geographic access to blood banking facilities, highlighting significant rural-urban and regional disparities. By integrating population distribution with travel time, we identify ‘blood deserts’. Future policies must focus on the equitable distribution of facilities based on travel times. Future research should focus on utilizing geospatial tools to optimize resource allocation and improve accessibility.

## Acknowledgments

We thank the Health Informatics Group, which manages e-Raktkosh, for sharing the list of blood banking facilities with us.

## Data Availability Statement

All data produced in the present study are available upon reasonable request to the authors.

## Ethics approval and consent to participate

Not applicable, as we did not use any human participants or animal subjects’ data. All data used in this manuscript were taken from public sources.

## Consent for publication

Not applicable

## Authors’ contributions

***Study concept and design:*** Siddhesh Zadey

***Acquisition, analysis, or interpretation of data:*** All authors

***Drafting of the manuscript:*** Gaurav Urs, Siddhesh Zadey

***Critical revision of the manuscript for important intellectual content:*** All authors

***Statistical analysis:*** Aamir Miyajiwala, Ankit Raj, Siddhesh Zadey

***Administrative, technical, or material support:*** Siddhesh Zadey

***Study supervision:*** Siddhesh Zadey

